# Opportunistic Upper Endoscopy at the Time of Screening Colonoscopy: Feasibility, Acceptability, and Patient Perspectives

**DOI:** 10.64898/2026.01.26.26344849

**Authors:** Haejin In, Katherine De la Torre Cisneros, Brijesh Rana, Priscille Myrthil-Harder, Alexandra Adams, Ishita Dalal, Anish Patel, Keerthana Kesavarapu, Zhongren Zhou, Nirag Jhala, Elizabeth Handorf, Anita Kinney

**Affiliations:** Division of Surgical Oncology, Rutgers Cancer Institute, New Brunswick, NJ; Department of Health Behavior, Society, and Policy, Rutgers School of Public Health, New Brunswick, NJ; Department of Gastroenterology, Rutgers Robert Wood Johnson Medical School, New Brunswick, NJ; Department of Pathology and Laboratory Medicine, Rutgers Cancer Institute, New Brunswick, NJ; Department of Biostatistics and Epidemiology, Rutgers Cancer Institute, New Brunswick, NJ

**Keywords:** Stomach Neoplasms, Endoscopy, Gastrointestinal, Mass Screening, Precancerous Conditions

## Abstract

**Objectives:** Gastric cancer (GC) is a leading cause of cancer mortality in the United States (U.S.), yet no routine screening strategy exists. Opportunistic upper endoscopy (EGD) performed during screening colonoscopy (EGD-SC) may provide a practical early detection approach. We evaluated the feasibility, acceptability, patient perspectives, and diagnostic yield of EGD-SC.

**Methods:** This single-center, open-label, single-arm prospective trial enrolled adults aged 45–80 years scheduled for colonoscopy without prior EGD in the past five years. Feasibility was assessed by enrollment, added procedural time, and safety. Acceptability, patient beliefs, motivators, barriers, and satisfaction were assessed using pre- and post-procedure surveys. Gastric biopsies evaluated for precancerous lesions.

**Results:** Of individuals contacted, 51.6% expressed interest and 26.6% enrolled (n=50; median age 56; 48% male; 68% high-risk). Median added time was 17 minutes (range 9–26), with no complications. All participants rated EGD-SC as satisfactory (100%) and 90% as acceptable; most preferred the combined procedure (97.5%) and would recommend it to family or friends (92.5%). Knowledge gaps were common: nearly half lacked awareness of GC risk factors; although 72% viewed screening as beneficial, only 23.3% perceived GC as severe, and none considered themselves highly susceptible. EGD found *H. pylori* infection (32%), atrophic gastritis (14%), and intestinal metaplasia (12%), with higher prevalence among high-risk participants.

**Conclusions:** EGD-SC is feasible, safe, and highly acceptable, with strong patient endorsement and meaningful detection of GC precursor lesions. These findings support risk-stratified EGD-SC as a promising and pragmatic strategy for GC prevention and early detection in the U.S.

## INTRODUCTION

Gastric Cancer (GC) is a highly lethal yet curable and preventable cancer ranked fifth in incidence and mortality worldwide.^1^ In the US, GC ranks fifteenth in incidence and accounts for approximately 30,000 new cases and 11,000 deaths annually.^2^ Despite being uncommon compared with other major cancers, GC remains a significant health concern, particularly among racial and ethnic minorities, immigrants from high-incidence regions, and individuals with a family history of GC. Strategies aimed at interception and early detection of GC in the US remain far less developed than those for other major cancers.^3^

Clinical and professional organizations have consistently underscored the need for GC screening guidelines. The most recent US guidelines, published by the American Gastroenterological Association (AGA) in 2025, represents an important step forward, but still provide no clear recommendations for GC screening, focusing instead on surveillance. Notably, the 2025 update expanded prior guidance by recommending three-year endoscopic surveillance for patients with intestinal metaplasia (IM) who additionally possess high-risk features—such as a family history of GC or belonging to a high-incidence racial or ethnic group.^4, 5^ This represents progress from the 2020 AGA guidelines, which advised against surveillance in patients with IM and limited its recommendations to high-risk histologic subtypes alone, while cautiously acknowledging that high-risk individuals could elect surveillance at their discretion. Despite this advancement, the current approach remains inherently inadequate, as it only benefits those who have already undergone esophagogastroduodenoscopy (EGD) and been identified with precursor lesions, leaving a large proportion of at-risk individuals unscreened. In contrast, national GC screening programs have been implemented in Japan since 1983 and in Korea since 2002,^6, 7^ resulting in substantial improvements in early detection and a 30-60% reduction in mortality.^8, 9^ The recently published 2025 European guidelines endorse population-based screening with EGD for individuals residing in intermediate- and high-risk regions.^10^

Conducting an EGD at the time of routine screening colonoscopy (EGD-SC) can integrate GC screening into existing preventive care workflows and may be a cost-efficient strategy to expand and promote GC screening, particularly among high-risk populations.^11, 12^ It is estimated that 63% of people aged 50-75 in the US undergo colorectal cancer screening with sigmoidoscopy or colonoscopy.^13^ Given the large number of patients presenting to endoscopy facilities for colorectal cancer screening, incorporating an EGD for GC screening during screening colonoscopy might be a highly practical and appealing approach.^14^

We report the findings of Pilot Study of <u>S</u>tomach <u>C</u>ance<u>R E</u>arly detection and prevention with <u>EN</u>doscopy (Pilot-SCREEN) (NCT05566899), in which we examined the feasibility, acceptability, satisfaction, and participant preference for EGD-SC screening. We also assessed GC screening perceptions, attitudes, and beliefs, as well as facilitators and barriers to GC screening. As an exploratory aim, we examined the diagnostic yield of EGD-SC and differences between high- and low-risk participants.

## METHODS

### Study design and Participants

Pilot-SCREEN study is a single-center, open-label, single-arm prospective clinical trial evaluating the addition of EGD at the time of a scheduled colonoscopy. Participants aged 45 to 80 years at the time of informed consent, who were scheduled for a colonoscopy but not an EGD, at a single academic institution, and willing and able to comply with all aspects of the protocol were enrolled. Exclusion criteria included the presence of any anatomic alteration that precludes EGD, a previous total gastrectomy, any medical condition that substantially increases the risk of EGD, or if the individual had undergone EGD in the last five years. Research staff contacted potential participants by phone to assess their willingness to participate and confirmed eligibility based on inclusion criteria. Reasons for non-participation were recorded.

Enrolled participants were contacted approximately 1 week prior to EGD to complete the pre-EGD survey. This survey assessed participants’ health history, beliefs, knowledge about GC, and motivators and barriers to undergoing an EGD, and was designed to take approximately 30 minutes to complete. Before EGD procedure, blood, urine, stool, and saliva were collected.

During the procedure, biopsy samples were taken from five sites in the stomach (lesser antrum, greater antrum, lesser body, greater body, and angular incisure) according to the Sydney Protocol.^15^ Procedure start and end times for both EGD and colonoscopy were recorded.

Procedure-related complications were monitored from the start of EGD until patient discharge and medical records were reviewed for any readmissions related to the EGD-colonoscopy procedure within one week of the procedure. A post-EGD survey was conducted 1-2 weeks after the procedure to evaluate participants’ preferences and satisfaction, requiring approximately 10 minutes to complete. Surveys were administered via email link, paper, or on a tablet provided by the research coordinator in the endoscopy suite, with English and Spanish versions available based on participant preference. This study received IRB approval (Pro2022001252).

For exploratory analyses, participants were classified as high or low risk for developing GC, with high risk defined as having a first-degree relative with GC, self-identifying as Hispanic, non-Hispanic Black, or Asian/Pacific Islander, or being a first-generation immigrant from regions with high GC incidence based on 2022 WHO incidence rates (ASR>10.3) such as Korea (38.4), Japan (40.9), Russia (19.8), China (19.5), and Central and South America (12.0-21.8).^16^

### Survey instruments

The pre-EGD survey collected information on sociodemographic characteristics, dietary and lifestyle factors, medical history, knowledge of GC risk factors, health beliefs, screening barriers perceived, provider recommendations, health literacy and language, risk assessment and screening practices, colonoscopy and endoscopy history, and cultural background. Participants’ perceptions of susceptibility, severity, benefits, efficacy, and barriers to GC screening were measured using validated items on a 5-point Likert scale (1=strongly disagree to 5=strongly agree).^17^ Composite scores were calculated for each domain.

The post-EGD survey captured participants’ attitudes toward the addition of EGD to the colonoscopy procedure, evaluating satisfaction, preferences, perceived efficacy, screening barriers, and EGD acceptability.^18, 19^ Assessment of EGD acceptability included items measuring affective attitude, procedure burden, ethical concerns, perceived effectiveness, intervention coherence, self-efficacy, opportunity cost, and overall acceptability.^18^ **Supplementary Table 1** lists all survey items and their sources from validated instruments. Further details on data collection and variable assessment are provided in the **Supplementary Methods**.

### Statistical Analysis

Descriptive statistics were calculated to determine the median and range for numerical variables, such as age and time. Frequency and percentages were computed for participants’ responses for each survey item. One-sample Wilcoxon signed-rank tests were calculated to evaluate the participants’ responses regarding the EGD procedure. The median values of the ordinal responses were calculated and compared to the neutral answer (neither agree nor disagree, no opinion or neutral) to assess the level of agreement with each question or statement. Additionally, the Mann-Whitney U test was used to examine the differences in survey responses between participants at high and low risk. All authors had access to the study data and reviewed and approved the final manuscript. The data were analyzed using SAS software version 9.4 (SAS Institute Inc., Cary, NC, USA).

## RESULTS

### Study Enrollment and Feasibility

A total of 369 patients aged ≥45 years who were scheduled for colonoscopy were identified through initial screening of the electronic health record. In a secondary chart review, 74 patients were excluded due to a prior EGD within five years. The remaining 295 patients were contacted by research staff for recruitment; among them, 188 (63.7%) were reached and approached for participation, while 107 (36.3%) could not be contacted due to lack of response. Among those contacted, 97 (51.6%) expressed interest in participating, while 91 (48.4%) did not. Interest in participation was higher among the high-risk group compared to the low-risk group (65.7% vs. 33.7%). Reasons for non-participation included rescheduling of colonoscopy to an unforeseen date, lack of interest in endoscopy, or fear of additional anesthesia (**Supplementary Table 2**). Among those who expressed interest, final eligibility was confirmed by telephone using the qualification checklist, resulting in 89 participants (91.8%) agreeing to enroll in the study, while 8 individuals were deemed ineligible. Of those who consented, 12 patients (13.5%) subsequently withdrew, and 27 patients (30.3%) did not complete the scheduled colonoscopy.

Ultimately, 50 participants underwent an EGD at the time of their colonoscopy, corresponding to 26.6% of all contacted patients (**Figure 1**). Pre-EGD survey was completed by 43 (86.0%) and post-EGD survey by 40 (80.0%) participants.

**Figure 1.**
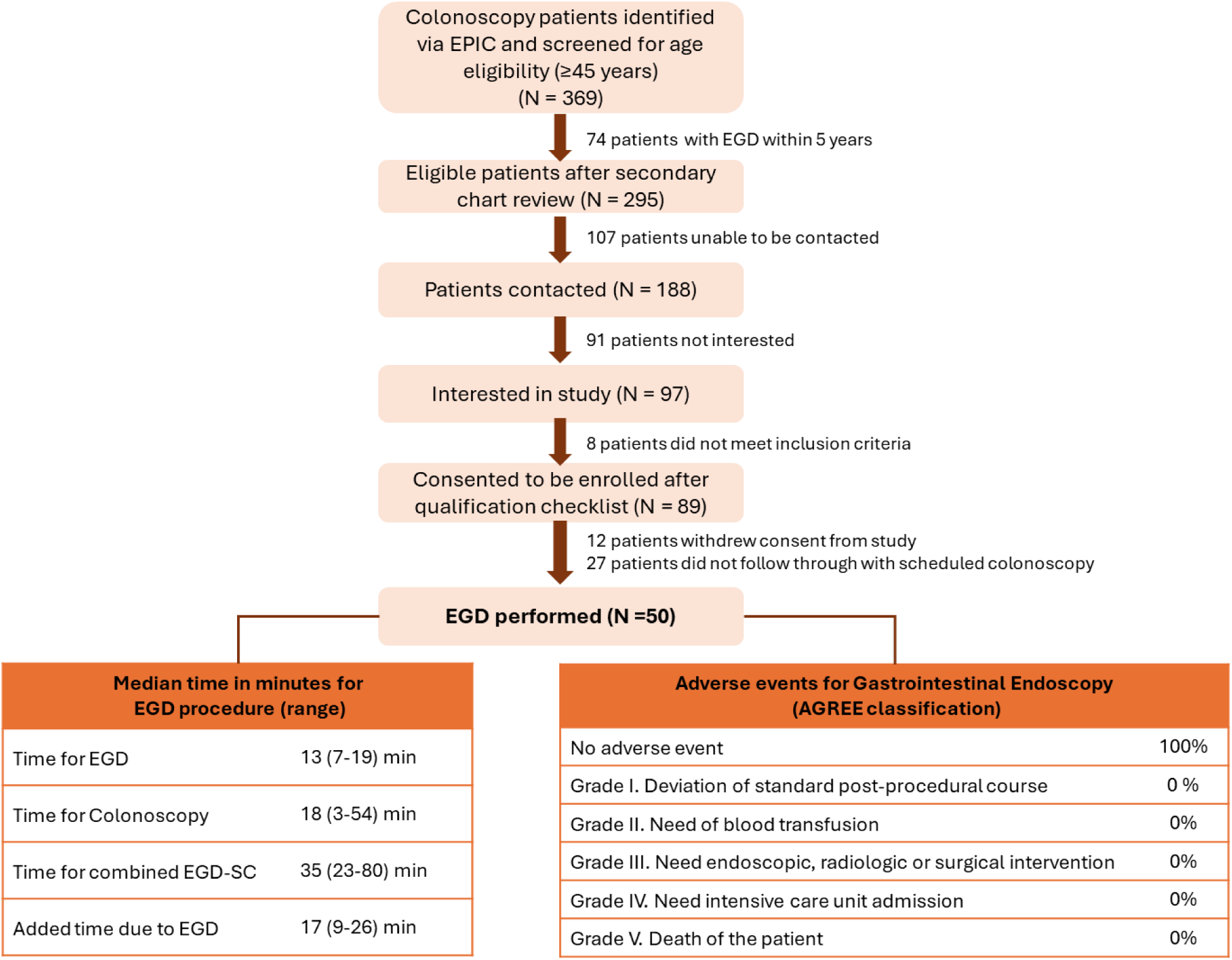
Enrollment Flow Chart.

### Study Population

**Table 1** summarizes the characteristics of the study population. The median age of enrolled participants was 56 (range 45-76), and 48% were male. Thirty-four percent were Non-Hispanic Whites (NHW), 22% non-Hispanic Black, 34% were Hispanics, and 8% were Asian. Overall, 46% of the participants were born in US, 24% born in countries with low GC incidence (e.g. Ghana, Guyana, Haiti, India, Italy, Jamaica, Nigeria, Pakistan, Senegal), and 30% born in high-risk countries (e.g. Colombia, Costa Rica, Dominican Republic, Ecuador, El Salvador, Guatemala, Mexico, Peru). Overall, 58.1% had a college education or higher, 32.6% had a high school education, and 9.3% had less than a high school. A family history of GC was reported in 6% of first-degree relatives. Approximately 50% had private insurance, 26% had Medicare or Medicaid, and 24% were uninsured; among them, 16.3% reported difficulty with transportation, and 53.5% reported difficulty understanding written medical information.

**Table 1.** Sociodemographic and clinical characteristics of participants.

|  | All sample<br>(n=50) | High-risk<br>(n=34) | Low-risk<br>(n=16) | p-value |
| --- | --- | --- | --- | --- |
| **Characteristics** | **N (%)** | **N (%)** | **N (%)** |  |
| Age (median, range) | 56 (45-76) | 54 (49-62) | 56 (45-76) | 0.2594 |
| **Sex** |  |  |  | 0.846 |
| Males | 24 (48.0) | 16 (47.1) | 8 (50.0) |  |
| Females | 26 (52.0) | 18 (52.9) | 8 (50.0) |  |
| **Race/Ethnicity** |  |  |  | <0.0001 |
| White, non-Hispanics | 17 (34.0) | 1 (2.9) | 16 (80.0) |  |
| Black, non-Hispanics | 11 (22.0) | 11 (32.4) | 0 (0.0) |  |
| Hispanics | 18 (36.0) | 18 (52.9) | 0 (0.0) |  |
| Asians | 4 (8.0) | 4 (11.8) | 0 (0.0) |  |
| **Place of birth/ immigration status?** |  |  |  | <0.0001 |
| US-born | 23 (46.0) | 8 (23.5) | 15 (93.8) |  |
| Foreign-born GC low-risk country | 12 (24.0) | 11 (32.4) | 1 (6.2) |  |
| Foreign-born GC high-risk country | 15 (30.0) | 15 (44.1) | 0 (0.0) |  |
| **Education level<sup>a</sup>** |  |  |  | 0.1919 |
| College or more | 25 (58.1) | 13 (50.0) | 12 (70.6) |  |
| High school | 14 (32.6) | 9 (34.6) | 5 (29.4) |  |
| Less than High school | 4 (9.3) | 4 (15.4) | 0 (0.0) |  |
| **Family history of GC** |  |  |  | 0.2752 |
| First-degree relative | 3 (6.0) | 3 (8.8) | 0 (0.0) |  |
| Second-degree relative | 3 (6.0) | 2 (5.9) | 1 (6.2) |  |
| No | 32 (64.0) | 19 (55.9) | 13 (81.3) |  |
| Unknown | 12 (24.0) | 10 (29.4) | 2 (12.5) |  |
| **Insurance type** |  |  |  | 0.0213 |
| Private | 25 (50.0) | 15 (44.1) | 10 (62.5) |  |
| Medicare | 7 (14.0) | 4 (11.8) | 3 (18.8) |  |
| Medicaid | 6 (12.0) | 3 (8.8) | 3 (18.8) |  |
| None | 12 (24.0) | 12 (35.3) | 0 (0.0) |  |
| **Lifestyle risk factors** |  |  |  |  |
| Current smoking | 5 (10.0) | 5 (14.7) | 0 (0.0) | 0.1313 |
| Current alcohol intake | 6 (12.0) | 4 (11.8) | 2 (12.5) | 0.3502 |
| **Comorbidities** |  |  |  |  |
| Diabetes mellitus | 19 (38.0) | 15 (44.1) | 4 (25.0) | 0.1939 |
| Hypertension | 23 (46.0) | 17 (50.0) | 6 (37.5) | 0.4081 |
| *H. pylori* infection history | 6 (12.0) | 5 (14.7) | 1 (6.3) | 0.8321 |
| Gastritis | 6 (12.0) | 5 (14.7) | 1 (6.3) | 0.2855 |
| Stomach ulcer | 3 (6.0) | 2 (5.9) | 1 (6.3) | 0.7863 |
| Inflammatory bowel disease | 3 (6.0) | 1 (2.9) | 2 (12.5) | 0.3151 |
| **Healthcare access<sup>a</sup>** |  |  |  |  |
| Transportation difficulty | 7 (16.3) | 6 (23.1) | 1 (5.9) | 0.0772 |
| Difficulty understanding written medical information | 23 (53.5) | 17 (65.4) | 6 (35.3) | 0.0677 |
| **GI symptoms in the past year<sup>a</sup>** |  |  |  | 0.8336 |
| Yes (symptomatic) | 27 (62.8) | 16 (61.5) | 11 (64.7) |  |
| No (asymptomatic) | 16 (37.2) | 10 (38.5) | 6 (35.3) |  |
| **Knowledge of GC risk factors<sup>a</sup>** |  |  |  | 0.7828 |
| Very knowledgeable | 0 (0.0) | 0 (0.0) | 0 (0.0) |  |
| Moderately knowledgeable | 8 (18.6) | 4 (15.4) | 4 (23.5) |  |
| Less knowledgeable | 15 (34.9) | 9 (34.6) | 6 (35.3) |  |
| Not knowledgeable | 20 (46.5) | 13 (50.0) | 7 (41.2) |  |
| Abbreviations: GC; gastric cancer; GI, gastrointestinal |  |  |  |  |
| <sup>a</sup> Based on 43 respondents who completed the pre-EGD survey |  |  |  |  |

Regarding health habits, 10% were current smokers, and 12% used alcohol daily or more. Diabetes affected 38% and hypertension 46%. In addition, 12% reported prior *Helicobacter pylori* (HP) infection, 12% had clinical gastritis, 6% had a history of stomach ulcer, and 6% had inflammatory bowel disease. Despite this study only enrolled participants who otherwise would not have undergone EGD, 62.8% reported gastrointestinal symptoms in the last year. Knowledge of GC risk factors, assessed using 20 items, with one point for each correct answer, was generally low. None of the participants were very knowledgeable (≥16 points, ≥80%), 18.6% were moderately knowledgeable (12–15, 60–79%), and nearly half of the participants (46.5%) were not knowledgeable at all (≤3, <40%).

Among all participants, 68% were classified as high-risk individuals. Median age was similar between groups (high-risk: 54 [49–62] years; low-risk: 56 [45–76] years), as was the proportion of males (47.1% vs. 50%). Compared to the low-risk group, high-risk individuals were less likely to have a college-level education or higher (50.0% vs. 70.0%) and more likely to have less than a high school education (15.4% vs. 0.0%). They were also less likely to have private insurance (44.1% vs. 62.5%) and more likely to be uninsured (35.0% vs. 0.0%; p-value = 0.0213). High-risk participants experienced greater barriers to health care, including transportation (23.1% vs. 5.9%) and understanding written medical information (65.4% vs. 35.3%). Reporting of gastrointestinal symptoms in the past year was similar in the high- and low-risk groups (61.5% vs. 64.7%). Knowledge of GC risk factors was lower in the high-risk group, with fewer participants being moderately knowledgeable (15.4% vs. 23.5%) and more being not knowledgeable (50.0% vs. 41%).

### Satisfaction, Preference and Acceptability of the EGD-SC

The post-EGD survey results on satisfaction, preference and acceptability of EGD at time of colonoscopy is presented in **Table 2**. Satisfaction with colonoscopy and EGD outcomes using 2 items. All respondents (100%) reported being satisfied with the EGD, and 95% found it easy to undergo EGD concurrently with colonoscopy.

**Table 2.**
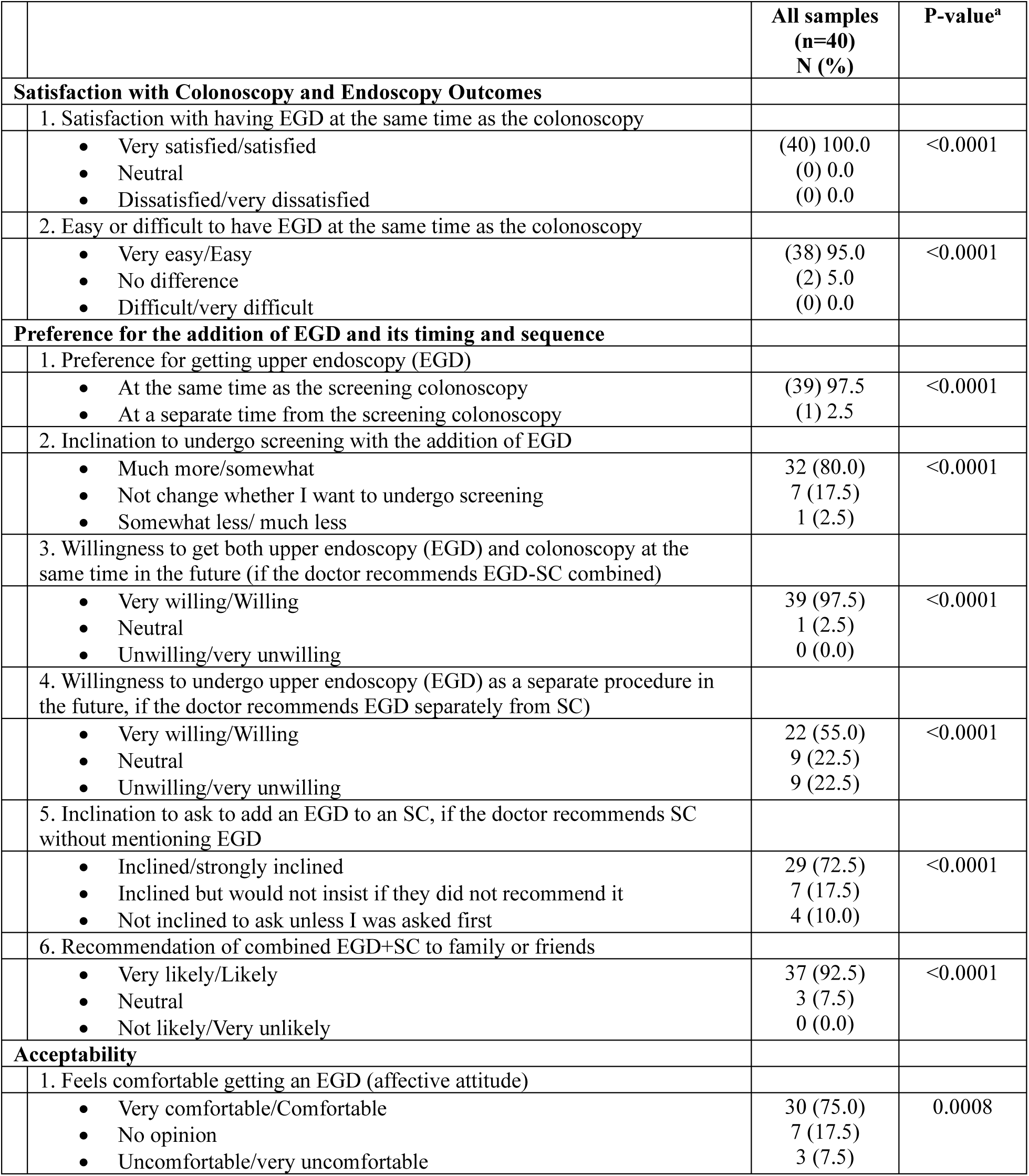
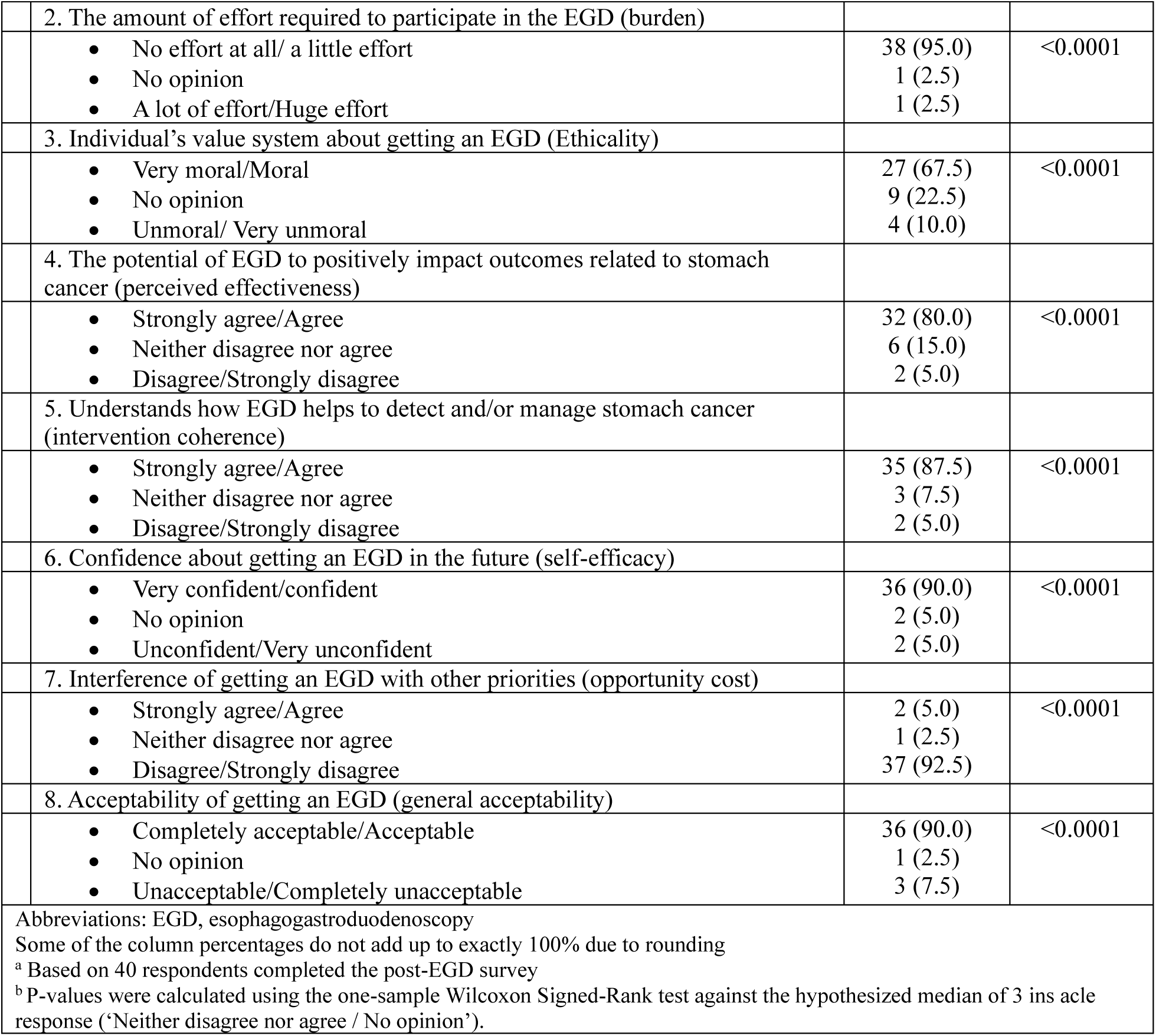
Post-survey on the perception of gastric cancer screening using a combined Endoscopy and Colonoscopy procedure.

|  |  | All samples<br>(n=40)<br>N (%) | P-value<sup>a</sup> |
| --- | --- | --- | --- |
| Satisfaction with Colonoscopy and Endoscopy Outcomes |  |  |  |
|  | 1. Satisfaction with having EGD at the same time as the colonoscopy |  |  |
|  | • Very satisfied/satisfied<br>• Neutral<br>• Dissatisfied/very dissatisfied | (40) 100.0<br>(0) 0.0<br>(0) 0.0 | <0.0001 |
|  | 2. Easy or difficult to have EGD at the same time as the colonoscopy |  |  |
|  | • Very easy/Easy<br>• No difference<br>• Difficult/very difficult | (38) 95.0<br>(2) 5.0<br>(0) 0.0 | <0.0001 |
| Preference for the addition of EGD and its timing and sequence |  |  |  |
|  | 1. Preference for getting upper endoscopy (EGD) |  |  |
|  | • At the same time as the screening colonoscopy<br>• At a separate time from the screening colonoscopy | (39) 97.5<br>(1) 2.5 | <0.0001 |
|  | 2. Inclination to undergo screening with the addition of EGD |  |  |
|  | • Much more/somewhat<br>• Not change whether I want to undergo screening<br>• Somewhat less/ much less | 32 (80.0)<br>7 (17.5)<br>1 (2.5) | <0.0001 |
|  | 3. Willingness to get both upper endoscopy (EGD) and colonoscopy at the same time in the future (if the doctor recommends EGD-SC combined) |  |  |
|  | • Very willing/Willing<br>• Neutral<br>• Unwilling/very unwilling | 39 (97.5)<br>1 (2.5)<br>0 (0.0) | <0.0001 |
|  | 4. Willingness to undergo upper endoscopy (EGD) as a separate procedure in the future, if the doctor recommends EGD separately from SC) |  |  |
|  | • Very willing/Willing<br>• Neutral<br>• Unwilling/very unwilling | 22 (55.0)<br>9 (22.5)<br>9 (22.5) | <0.0001 |
|  | 5. Inclination to ask to add an EGD to an SC, if the doctor recommends SC without mentioning EGD |  |  |
|  | • Inclined/strongly inclined<br>• Inclined but would not insist if they did not recommend it<br>• Not inclined to ask unless I was asked first | 29 (72.5)<br>7 (17.5)<br>4 (10.0) | <0.0001 |
|  | 6. Recommendation of combined EGD+SC to family or friends |  |  |
|  | • Very likely/Likely<br>• Neutral<br>• Not likely/Very unlikely | 37 (92.5)<br>3 (7.5)<br>0 (0.0) | <0.0001 |
| Acceptability |  |  |  |
|  | 1. Feels comfortable getting an EGD (affective attitude) |  |  |
|  | • Very comfortable/Comfortable<br>• No opinion<br>• Uncomfortable/very uncomfortable | 30 (75.0)<br>7 (17.5)<br>3 (7.5) | 0.0008 |

|  |  |  |
| --- | --- | --- |
| 2. The amount of effort required to participate in the EGD (burden) |  |  |
| No effort at all/ a little effort<br>No opinion<br>A lot of effort/Huge effort | 38 (95.0)<br>1 (2.5)<br>1 (2.5) | <0.0001 |
| 3. Individual's value system about getting an EGD (Ethicality) |  |  |
| Very moral/Moral<br>No opinion<br>Unmoral/ Very unmoral | 27 (67.5)<br>9 (22.5)<br>4 (10.0) | <0.0001 |
| 4. The potential of EGD to positively impact outcomes related to stomach cancer (perceived effectiveness) |  |  |
| Strongly agree/Agree<br>Neither disagree nor agree<br>Disagree/Strongly disagree | 32 (80.0)<br>6 (15.0)<br>2 (5.0) | <0.0001 |
| 5. Understands how EGD helps to detect and/or manage stomach cancer (intervention coherence) |  |  |
| Strongly agree/Agree<br>Neither disagree nor agree<br>Disagree/Strongly disagree | 35 (87.5)<br>3 (7.5)<br>2 (5.0) | <0.0001 |
| 6. Confidence about getting an EGD in the future (self-efficacy) |  |  |
| Very confident/confident<br>No opinion<br>Unconfident/Very unconfident | 36 (90.0)<br>2 (5.0)<br>2 (5.0) | <0.0001 |
| 7. Interference of getting an EGD with other priorities (opportunity cost) |  |  |
| Strongly agree/Agree<br>Neither disagree nor agree<br>Disagree/Strongly disagree | 2 (5.0)<br>1 (2.5)<br>37 (92.5) | <0.0001 |
| 8. Acceptability of getting an EGD (general acceptability) |  |  |
| Completely acceptable/Acceptable<br>No opinion<br>Unacceptable/Completely unacceptable | 36 (90.0)<br>1 (2.5)<br>3 (7.5) | <0.0001 |
| Abbreviations: EGD, esophagogastroduodenoscopy<br>Some of the column percentages do not add up to exactly 100% due to rounding<br><sup>a</sup> Based on 40 respondents completed the post-EGD survey<br><sup>b</sup> P-values were calculated using the one-sample Wilcoxon Signed-Rank test against the hypothesized median of 3 ins acle response ('Neither disagree nor agree / No opinion'). |  |  |

Preferences regarding the addition of EGD and its timing and sequence were assessed using 8 items. Overall, participants demonstrated a strong preference for combining EGD with colonoscopy. Specifically, 97.5% of participants reported they would be willing to undergo both procedures if recommended by their physician, and an equal proportion (97.5%) preferred having the EGD performed at the same time as the colonoscopy. 90% of participants indicated they would request the addition of EGD if colonoscopy were recommended without mention of EGD, with 72.5% expressing a strong intention to pursue this request. The addition of EGD was also perceived as an incentive to undergo screening: 80.0% of participants reported they would be more likely to undergo screening if EGD were included alongside colonoscopy. If EGD were recommended as a separate procedure, 55% indicated they would be very willing to undergo it, whereas 22.5% reported they would be unwilling. Finally, nearly all participants (92.5%) stated they would recommend the combined EGD and colonoscopy procedure to a family member or friend.

Nearly all (90%) participants considered the combined EGD-SC procedure acceptable. A total of 95.0% reported little or no effort was required to participate, 90.0% felt confident about undergoing EGD in the future, 87.5% understood how EGD could help detect and/or manage stomach cancer, and 80% believed that EGD has the potential to positively impact stomach cancer outcomes. Additionally, 75.0% reported feeling comfortable with the procedure, 17.5% indicated neither comfort nor discomfort, and 7.5% (n=3) reported discomfort. Only one report of discomfort was directly related to the procedure (a mild sore throat after the procedure), whereas the remaining two reports were related to administrative factors, specifically long wait times on the day of the procedure. Furthermore, 67.5% believed that undergoing EGD is moral and 5% indicated that getting an EGD interfered with other priorities.

Although overall acceptability of combined EGD with screening colonoscopy did not differ significantly between high- and low-risk groups, notable differences emerged. High-risk participants were less likely to feel comfortable with EGD (70.4% vs. 84.6%), believe it could positively impact GC outcomes (70.4% vs. 100.0%), and report it not interfering with future priorities (88.9% vs. 100.0%). The acceptability of the combined procedure remained high in both groups (81.5% vs. 92.3%). High-risk participants were also more likely to express concerns related to confidence, ease, effectiveness, and interference with priorities (50% vs. 0%) (**Supplementary Table 3**). These findings indicate that while EGD is generally acceptable, high-risk individuals face greater procedural and logistical barriers that should be considered in targeted screening strategies.

Participants’ perceptions of EGD efficacy and barriers to screening changed modestly after completing the combined EGD–screening colonoscopy procedure (**Table 3).** Agreement that EGD is easy to undergo remained stable (39.5% pre vs. 37.5% post), but the proportion who disagreed declined substantially (44.2% to 15.0%), with many shifting to neutral responses (16.3% to 42.5%). Knowledge that EGD provides information about stomach cancer was high at baseline and showed minimal change (83.7% to 85.0%). Perceived convenience also shifted: disagreement dropped sharply (48.8% to 2.5%), neutral responses increased (4.7% to 47.5%), and agreement remained stable. Overall, the procedure reduced negative perceptions of EGD efficacy, leading participants to adopt more neutral evaluations. Perceived barriers to screening also shifted toward reduced concern, with increases in the proportion disagreeing that they had too many competing priorities (37.2% to 80.0%) or that GC screening was a low priority (24.9% to 85.0%). Perceptions related to caregiving responsibilities showed little change (83.7% to 82.5%).

**Table 3.**
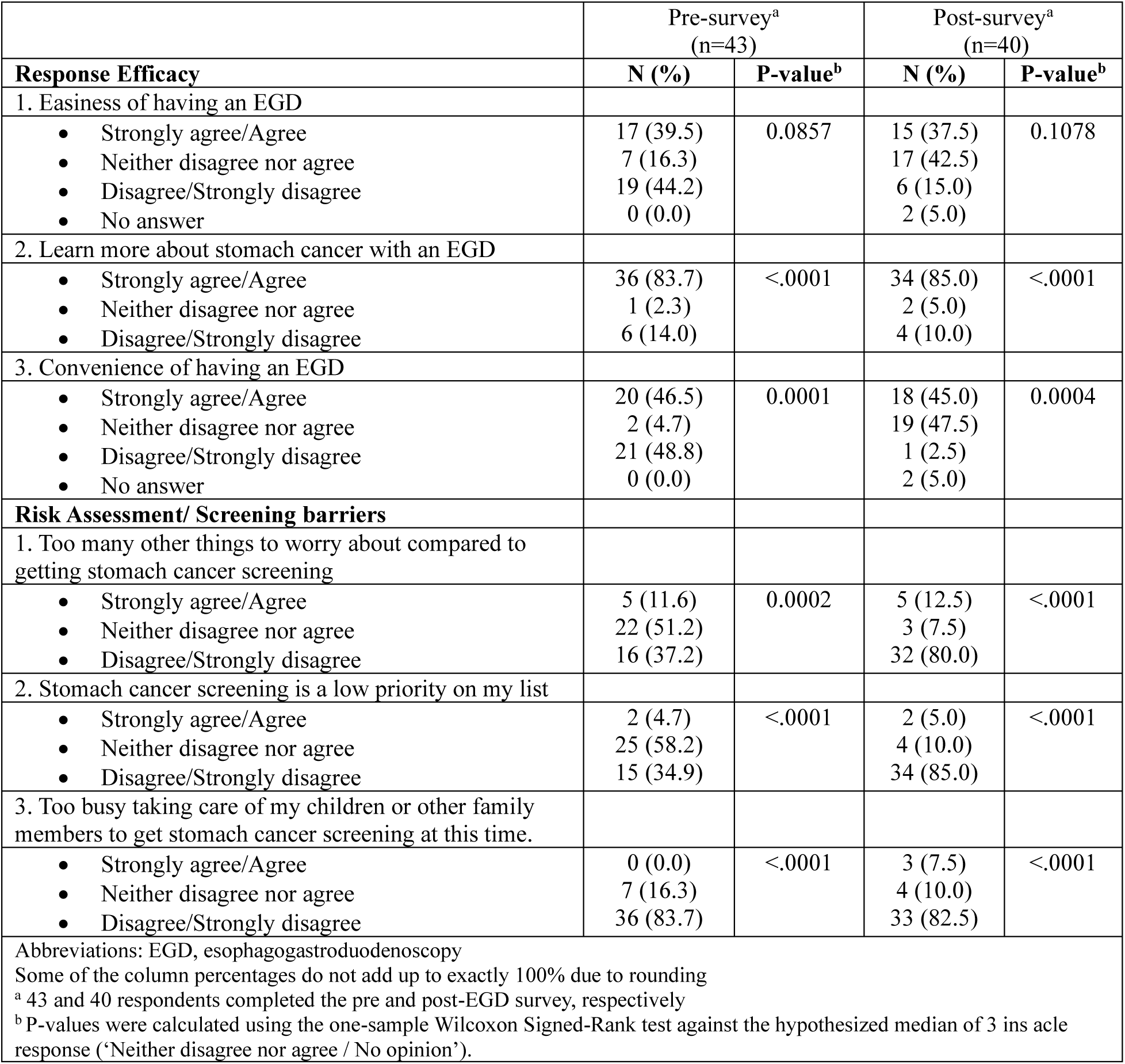
Perceptions about EGD efficacy and risk assessment before and after combined EGD-SC procedure.

|  | Pre-survey<sup>a</sup><br>(n=43) |  | Post-survey<sup>a</sup><br>(n=40) |  |
| --- | --- | --- | --- | --- |
| **Response Efficacy** | **N (%)** | **P-value<sup>b</sup>** | **N (%)** | **P-value<sup>b</sup>** |
| 1. Easiness of having an EGD |  |  |  |  |
| Strongly agree/Agree<br>Neither disagree nor agree<br>Disagree/Strongly disagree<br>No answer | 17 (39.5)<br>7 (16.3)<br>19 (44.2)<br>0 (0.0) | 0.0857 | 15 (37.5)<br>17 (42.5)<br>6 (15.0)<br>2 (5.0) | 0.1078 |
| 2. Learn more about stomach cancer with an EGD |  |  |  |  |
| Strongly agree/Agree<br>Neither disagree nor agree<br>Disagree/Strongly disagree | 36 (83.7)<br>1 (2.3)<br>6 (14.0) | <.0001 | 34 (85.0)<br>2 (5.0)<br>4 (10.0) | <.0001 |
| 3. Convenience of having an EGD |  |  |  |  |
| Strongly agree/Agree<br>Neither disagree nor agree<br>Disagree/Strongly disagree<br>No answer | 20 (46.5)<br>2 (4.7)<br>21 (48.8)<br>0 (0.0) | 0.0001 | 18 (45.0)<br>19 (47.5)<br>1 (2.5)<br>2 (5.0) | 0.0004 |
| **Risk Assessment/ Screening barriers** |  |  |  |  |
| 1. Too many other things to worry about compared to getting stomach cancer screening |  |  |  |  |
| Strongly agree/Agree<br>Neither disagree nor agree<br>Disagree/Strongly disagree | 5 (11.6)<br>22 (51.2)<br>16 (37.2) | 0.0002 | 5 (12.5)<br>3 (7.5)<br>32 (80.0) | <.0001 |
| 2. Stomach cancer screening is a low priority on my list |  |  |  |  |
| Strongly agree/Agree<br>Neither disagree nor agree<br>Disagree/Strongly disagree | 2 (4.7)<br>25 (58.2)<br>15 (34.9) | <.0001 | 2 (5.0)<br>4 (10.0)<br>34 (85.0) | <.0001 |
| 3. Too busy taking care of my children or other family members to get stomach cancer screening at this time. |  |  |  |  |
| Strongly agree/Agree<br>Neither disagree nor agree<br>Disagree/Strongly disagree | 0 (0.0)<br>7 (16.3)<br>36 (83.7) | <.0001 | 3 (7.5)<br>4 (10.0)<br>33 (82.5) | <.0001 |
| Abbreviations: EGD, esophagogastrroduodenoscopy<br>Some of the column percentages do not add up to exactly 100% due to rounding<br><sup>a</sup> 43 and 40 respondents completed the pre and post-EGD survey, respectively<br><sup>b</sup> P-values were calculated using the one-sample Wilcoxon Signed-Rank test against the hypothesized median of 3 ins acle response ('Neither disagree nor agree / No opinion'). |  |  |  |  |

### Complications and added time to Colonoscopy due to Endoscopy

No enrolled participants experienced adverse events requiring extended observation after the procedure. There were no deviations from the standard post-procedural care, and no participants required pharmacologic, endoscopic, radiological, or surgical intervention, hospital admission, or experienced mortality. The median additional time required for the procedure due to EGD was 17 minutes (range 9-26), with a total median time of 35 minutes (range 23-80) for the combined EGD and colonoscopy. This time frame included patient positioning, the EGD procedure itself, and the interval before the start of the colonoscopy. The longest durations were observed in patients with clinically evident lesions in the endoscopic examination. Each EGD procedure (from endoscope intubation to extubation time) had a median of 13 minutes, with a minimum of 7 minutes (**Figure 1**).

### Baseline Screening Behaviors

Of respondents, 55.8% reported having undergone a previous colonoscopy, and 88% considered colonoscopy to be critical for personal health and well-being. The primary reason for scheduling their current colonoscopy was a provider recommendation (63.8%). Reported reasons for the procedure included colon cancer screening (58.1%), follow-up of previously identified polyps (20.9%), screening for non-cancerous diseases (14.0%), a diagnostic evaluation for gastrointestinal disease symptoms (4.7%) and follow-up for prior positive test result (2.3%). The main reasons for choosing colonoscopy screening over other screening options (i.e. fecal test ) were a physician’s recommendation (60.5%), the belief that colonoscopy is better suited for screening and diagnosis (41.9%), perception of its importance (41.9%), reduction of cancer-related anxiety (30.2%), greater reliability compared to fecal testing (30.2%), and insurance coverage (23.3%).

When asked about GC screening, 19.5% of respondents reported that a healthcare provider had previously recommended GC screening. Only 14% had previously discussed their personal risk for GC with their provider. (**Supplementary Table 4**).

### Gastric Cancer Screening Perceptions, Attitudes, and Beliefs

Perceptions of GC susceptibility and severity, screening barriers and benefits were assessed using 38 items (**Table 4**). Overall, participants did not perceive themselves as being susceptible to GC, with 32.6% disagreeing or strongly disagreeing with statements reflecting their perceived likelihood of developing cancer and 67.4% with neutral responses. Most participants (74.4%) expressed neutral views on the severity of GC, while only 23.3% considered it a serious and harmful disease. Perceived barriers to screening were low, with most participants disagreeing (55.8%) or remaining neutral (44.5%) when asked about barriers to obtaining screening. Among barriers, the most agreed-upon concerns were fear of discovering an abnormality (25.6%) and cost (18.6%). In contrast, perceived benefits of screening benefits received the highest overall agreement (72.1%), particularly for statements such as “screening is a way to take care of myself” (97.7%), “GC can be found early with screening” (93.0%), and “the best way to find smaller cancers is by undergoing screening” (93%).

**Table 4.**
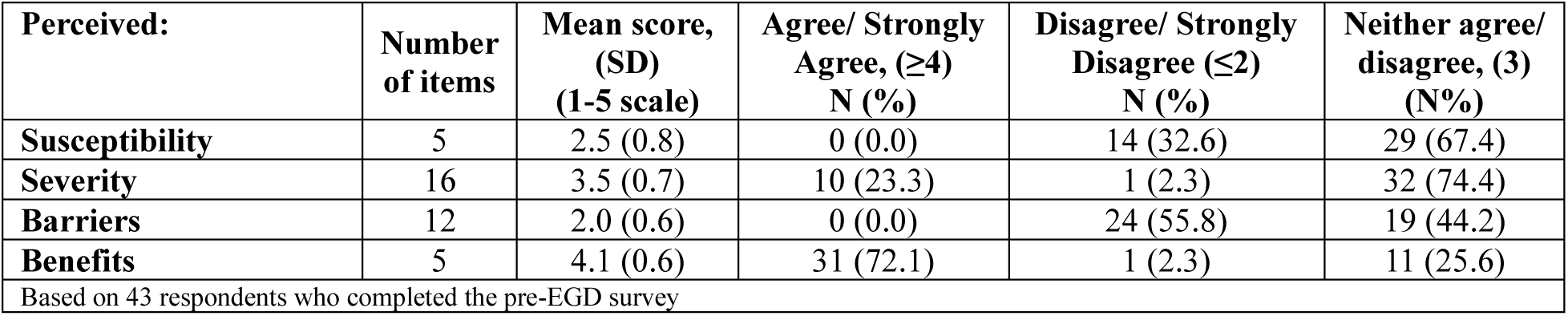
Perceptions and beliefs of threat, benefit and barriers about gastric cancer screening.

| Perceived: | Number of items | Mean score, (SD) (1-5 scale) | Agree/ Strongly Agree, ($\geq 4$) N (%) | Disagree/ Strongly Disagree ($\leq 2$) N (%) | Neither agree/ disagree, (3) (N%) |
| --- | --- | --- | --- | --- | --- |
| **Susceptibility** | 5 | 2.5 (0.8) | 0 (0.0) | 14 (32.6) | 29 (67.4) |
| **Severity** | 16 | 3.5 (0.7) | 10 (23.3) | 1 (2.3) | 32 (74.4) |
| **Barriers** | 12 | 2.0 (0.6) | 0 (0.0) | 24 (55.8) | 19 (44.2) |
| **Benefits** | 5 | 4.1 (0.6) | 31 (72.1) | 1 (2.3) | 11 (25.6) |
| Based on 43 respondents who completed the pre-EGD survey |  |  |  |  |  |

No statistically significant differences were observed between high-risk and low-risk groups in perceived threat, benefits, or barriers to GC screening (**Supplementary Table 5**). High-risk participants disagreed less often about there being barriers to screening than low-risk participants (44.4% vs. 75.0%); and were more inclined to select neutral responses. Similarly, the high-risk group perceived fewer benefits from screening compared to the low-risk participants (66.7% vs. 81.3%). In specific items, high-risk participants were more likely to agree that the perceived risk as *“GC is a hopeless disease”* (29.6% vs. 0.0%) while lower levels of agreement were observed among high-risk participants for the perceived benefits *“GC can be found early by undergoing screening”* (88.9% vs. 100.0%) and *“screening is a way to take care of myself”* (96.3% vs. 100.0%).

### Screening Intention after EGD

This item assessed participants’ willingness to engage in future conversations about GC screening. Before undergoing EGD, 32.6% of participants reported that they would definitely discuss EGD-based GC screening with their clinician within the next six months; this proportion increased to 46.3% after the procedure. Consistent with this finding, we also observed a modest overall decrease in expressed hesitancy toward considering GC screening. (**Supplementary Table 6**).

### Diagnostic Yield of Opportunistic EGD

Among participants, none of whom would have otherwise undergone EGD, biopsy specimens showed that 32% (95% CI 19.1-44.9) had active HP infection, 14% (4.4-23.6) had atrophic gastritis (AG), and 12% (3.0-21.0) had IM. No cases of dysplasia or neoplasia were identified. Overall, 18% (95% CI 7.4-28.7) of participants had had at least one abnormality (AG, IM, or dysplasia), and 40% (26.4-53.6) had at least HP or one abnormality. Findings were more pronounced in high-risk participants, who had a higher prevalence of HP (35.5% vs. 25%), AG (20.6% vs. 0.0%), and IM (14.7% vs. 6.3%) compared with low-risk individuals. At the individual level, 23.5% of high-risk participants had at least one condition (AG, IM, or dysplasia) compared with 6.3% of low-risk participants **(Table 5).** The prevalence of premalignant gastric lesions and composite pathology outcomes did not differ between participants with and without gastrointestinal symptoms (P>0.1 for all pathological findings) (**Supplementary Table 7**)

**Table 5.** Pathologic findings identified by opportunistic EGD according to gastric cancer risk.

|  | Total sample (n=50) | High-risk (n=34) | Low-risk (n=16) |
| --- | --- | --- | --- |
| **Pathology results** |  |  |  |
| *H. pylori* (+) | 16 (32.0) | 12 (35.3) | 4 (25.0) |
| Neutrophils (+) | 17 (34.0) | 13 (38.2) | 4 (25.0) |
| Monocyte (+) | 43 (86.0) | 32 (94.12) | 11 (68.8) |
| Atrophy (+) | 7 (14.0) | 7 (20.6) | 0 (0.0) |
| Intestinal Metaplasia (+) | 6 (12.0) | 5 (14.7) | 1 (6.3) |
| Dysplasia(+) | 0 (0.0) | 0 (0.0) | 0 (0.0) |
| **Individual level combined pathology** |  |  |  |
| HP, AG, IM or dysplasia | 20 (40.0) | 16 (47.1) | 4 (25.0) |
| AG, IM or dysplasia | 9 (18.0) | 8 (23.5) | 1 (6.3) |
| Abbreviations: HP, Helicobacter pylori; AG, atrophic gastritis; IM, intestinal metaplasia |  |  |  |

### Differences Between Routine Clinical EGD Report vs. Screening Study Report

**Supplementary Table 8** summarizes the clinically reported findings for the study participants. On clinical endoscopy reports, 20% were reported to have gastric polyps and 66% had gastritis. Esophageal findings were minimal, with mild and severe esophagitis reported in 15% and 2% of individuals, respectively. Gastric biopsies were obtained in 62% of participants and esophageal biopsies in 4% as part of standard clinical care.

Compared to routine clinical findings, research biopsies obtained via the Sydney Protocol identified AG (2% vs. 14%, p = 0.001), IM (12% vs. 4%, p = 0.007), and HP (32% vs. 24%, p = 0.003) at significantly higher frequencies **(Figure 2).**

**Figure 2.**
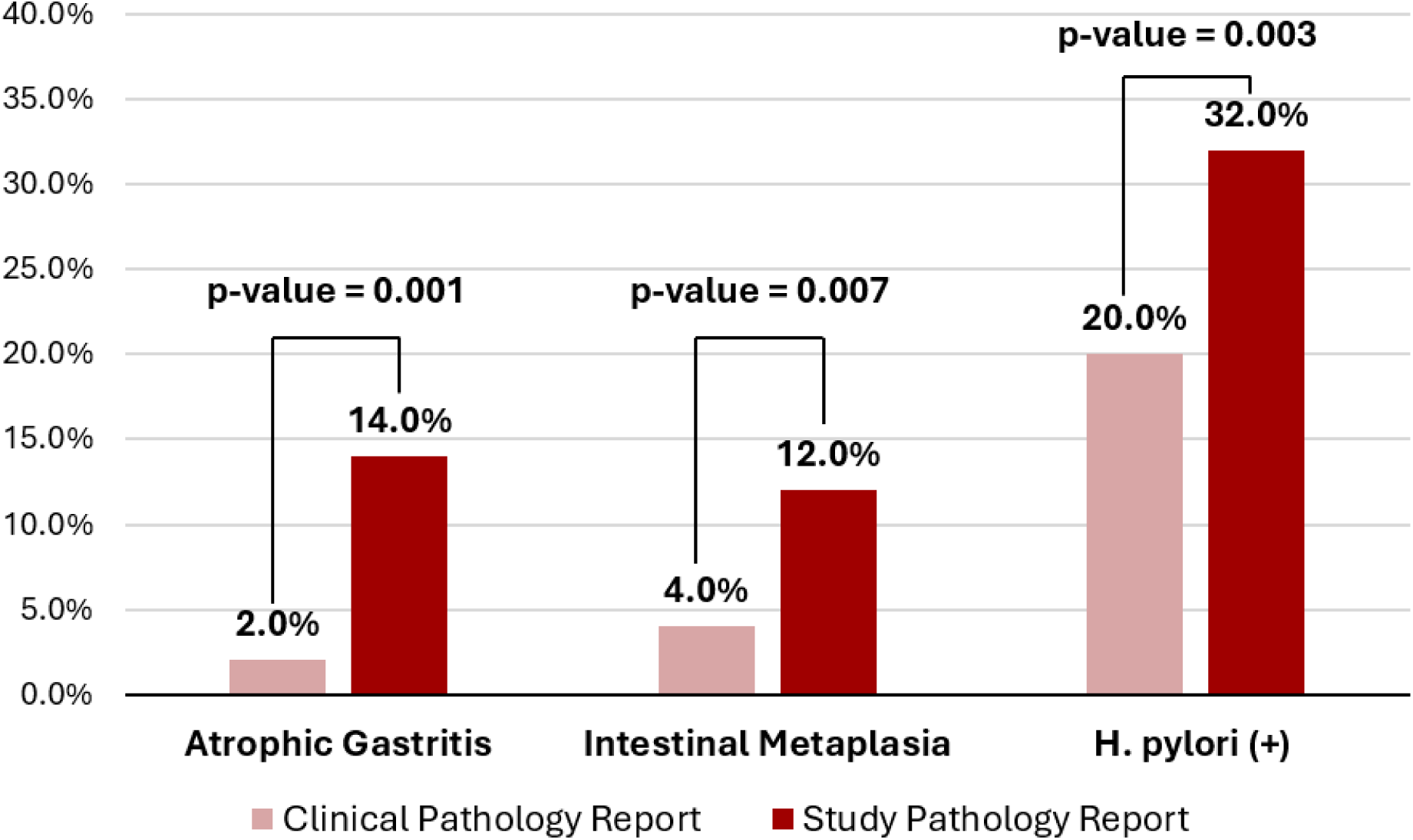
Differences between clinical routine report and screening report using Sydney protocol.

## DISCUSSION

Our study demonstrates that including an esophagogastroduodenoscopy at the time of screening colonoscopy (EGD-SC) is both feasible and acceptable. Participants reported high levels of satisfaction and preference for the combined procedure, and most considered GC screening to be valuable and EGD to be an effective method for early detection. In addition, this study provides valuable insights into perceptions of threat, benefit, and barriers to GC screening, as well as screening behaviors. Collectively, these findings support further investigation of EGD-SC as a potentially viable strategy for GC screening in high-risk U.S. populations, with the critical next step being evaluation of whether this approach leads to earlier-stage detection and, ultimately, reductions in GC-related mortality.

More than half of the individuals invited to participate expressed interest in receiving an EGD during their scheduled colonoscopy (51.6%). This level of interest closely aligns with findings from a study of U.S. veterans, in which 50.8% of eligible participants similarly indicated willingness to undergo a combined procedure.^20^ In Portugal, an intermediate-risk country, the proportion of patients agreeing to undergoing EGD along with colonoscopy was notably higher, at 94.8%,^21^ possibly due to the proactive, patient-centered approach taken by the gastroenterology team, which provided clear and personalized information about the potential benefits of adding EGD to the screening visit.

Reluctance to participate in combined endoscopy likely reflects broader gaps in awareness about GC and its prevention in the U.S. Unlike breast or colorectal cancer (CRC),^22, 23^ which benefits from well-established screening programs and strong provider endorsement, GC remains a low-priority condition for most patients. Concerns about anesthesia and scheduling conflicts may serve as proxies for deeper barriers, such as fear of complications or perceived inconvenience. Our findings of screening behaviors underscore a critical gap in awareness and provider engagement around GC prevention. While colorectal cancer screening is widely normalized through strong clinical and public health endorsement, GC screening remains largely absent from routine discussions. This lack of visibility likely contributes to low patient awareness, uncertainty about the value of EGD, and limited perceived relevance. Encouragingly, participants expressed strong willingness to undergo combined endoscopy when recommended by their physician, underscoring provider guidance as a key driver of uptake. These results emphasize the need for systematic integration of GC screening into preventive care, supported by targeted education and consistent provider recommendations. Improving knowledge of GC risk factors and normalizing EGD as part of standard practice will be essential to increase screening participation. Post-EGD survey findings demonstrated overwhelmingly positive perceptions of the combined EGD-SC procedure, highlighting that EGD-SC is not only feasible but also acceptable to patients. The combined approach was perceived as convenient and aligned with the goals of preventive care. Importantly, the few concerns raised were minor and largely related to administrative or logistical issues rather than the procedure itself, suggesting that operational refinements could further improve implementation. For the procedure itself, our study found that the median additional time required to include EGD was 17 minutes (range: 9–26). This modest additional time did not impose a significant burden on patients or the endoscopy team. These findings support the practicality of incorporating EGD into routine screening colonoscopy workflows and highlight its potential to increase GC screening uptake among populations that might otherwise remain unscreened.

Our study found that while some participants viewed GC as a serious disease, the majority did not perceive themselves to be highly susceptible to developing GC. In contrast, studies of CRC screening have reported high perceived susceptibility to CRC, likely reflecting greater public awareness of CRC as a common disease in the US.^24^ While the perceived benefits of CRC screening were similarly high, the nature of the barriers differs between the two cancers. For CRC, uncertainty about scheduling and fear of pain were mostly cited, whereas for GC, concerns centered on cost and fear of discovering abnormal findings. In the high-risk group, lower perceived benefits of screening, higher perceived barriers, limited knowledge of GC risk factors, transportation challenges, and difficulty understanding medical information should be considered when designing interventions. These differences suggest that awareness of GC remains low in the US and that targeted educational efforts, in addition to policy changes, may be necessary to overcome both emotional and financial barriers to screening.

GC screening with EGD provides three distinct opportunities to decrease GC morbidity and mortality. First, it has the potential to discover cancer in early stages before the onset of symptoms, leading to higher rates of survival. Second, it enables the identification and removal of precancerous lesions, thereby preventing the development of cancer. Third, EGD facilitates the detection and treatment of active HP infection, a primary risk factor for GC, providing an opportunity for primary prevention of GC. HP infection drives the majority of GC, accounting for 75–89% of cases.^25^ As the most common chronic bacterial infection worldwide, HP prevalence varies by geography, birth cohort, and race/ethnicity in the United States.^26^ Chronic infection initiates the “Correa cascade,” progressing from chronic gastritis to AG, IM, dysplasia, and ultimately adenocarcinoma.^27^ Yet, population-level prevalence of GPMC among asymptomatic individuals or those who would not otherwise undergo EGD is largely unknown, as most data derive from symptomatic patients presenting with dyspepsia/abdominal pain, weight loss, or gastrointestinal bleeding. Cross-sectional studies of clinically indicated endoscopies indicate higher prevalence among Asians (26–30%) and Hispanics (12–14%) compared with NHW (8–9%).^28, 29^ However, importantly, the presence of symptoms poorly correlates with GPMC,^30^ underscoring the inadequacy of symptom-based detection. Reliance on clinical presentation alone risks missed opportunities for early identification, prevention, and curative treatment of GC.

Despite the small sample size, opportunistic EGD detected gastric premalignant conditions (GPMC) in 18% of participants, which significantly exceeded the detection rates by routine clinical assessment. This finding is consistent with a study in U.S. veterans, where research EGD performed during colonoscopy identified IM at higher rates in asymptomatic individuals (21.4%) than in symptomatic patients seen in gastrointestinal clinics (18.3%).^20^ Further research is warranted to investigate the prevalence of GPMC, explore barriers to and motivators for combined EGD-SC in high-risk U.S. populations, and assess the potential impact of screening on reducing GC mortality.

EGD-SC strategy represents a viable and efficient option for integrating GC screening into established colorectal cancer (CRC) screening programs. Performing both EGD and colonoscopy during the same session allows simultaneous evaluation of the oropharynx, esophagus, stomach, colon, rectum and anus, thereby expanding the preventive reach of a single screening encounter. This combined approach minimizes patient burden, reduces procedural redundancy, and leverages existing endoscopic infrastructure to enhance the efficiency and comprehensiveness of cancer prevention. Modeling studies of US populations suggest that EGD-SC is a promising strategy for reducing GC mortality in the US, especially for high-risk populations.^11, 14^

The study has limitations. The single-institution design may limit generalizability, as acceptability and feasibility may vary across clinical settings; however, the sample size was sufficient to provide stable estimates for the primary outcomes. The reliance on self-reported data introduces potential response bias. Exploratory analyses were constrained by limited power and incomplete pre- and post-EGD survey data, and pathologic findings based on small sample sizes should be interpreted with caution. Future multicenter studies with larger, more diverse populations are needed to confirm generalizability and further explore subgroup and regional variation.

### Conclusions

Performing EGD at the time of screening colonoscopy is a feasible and well-received strategy for gastric cancer screening. The combined EGD-SC approach is widely perceived as satisfactory and preferred. Participants recognized the value of GC screening and viewed EGD as an effective method for early detection. However, targeted strategies are needed to improve the accuracy of patients’ self-perceived susceptibility and to reduce barriers among high-risk populations. Expanding public awareness, strengthening provider engagement, and establishing policy-level recommendations for high-risk groups will be critical to advancing GC screening efforts in the United States.

## Data Availability

All data produced in the present study are available upon reasonable request to the authors.

## Data availability statement

Data will be made available to other researchers upon reasonable request.

