## Supplemental material for "Opportunistic Upper Endoscopy at the Time of Screening Colonoscopy: Feasibility, Acceptability, and Patient Perspectives"

**Supplementary Material**

### Supplementary Methods

#### Data Collection and Variable Assessment

Demographic and clinical variables, including age, sex, race/ethnicity, insurance type, comorbidities, tobacco and alcohol use, were extracted from clinical records.

The pre-EGD survey assessed dietary habits, lifestyle, medical history, current gastrointestinal symptoms, knowledge of gastric cancer (GC) risk factors, health beliefs, screening barriers, perceived risk, provider recommendations, health literacy, language, risk assessment, and cultural background (Supplementary Table 1). From the pre-survey, education was categorized as high (some college, ≥10 years of college credit without a degree, bachelor's, master's, professional, or doctoral degree) or low (no schooling, kindergarten, 12th grade–no diploma, high school diploma, GED, or equivalent). GC risk factor knowledge was evaluated using 20 items (e.g., diet, alcohol, smoking, stress, obesity, physical inactivity, Helicobacter pylori infection, gastric ulcer, chronic gastritis, prior stomach surgery, family history, female sex, and birth outside the US). Responses were scored as correct (1 point) or incorrect/do not know. Knowledge was categorized as very knowledgeable (≥16 points, ≥80%), moderately knowledgeable (12–15 points, 60–79%), less knowledgeable (4–11 points, 40–59%), and not knowledgeable (≤3 points, <40%). Participants reported gastrointestinal symptoms and frequency over the past year. Symptomatic individuals were defined as those who reported experiencing stomach pain or nausea at least once per week; vomiting at least once per month; frequent bloating; changes in appetite; or interruptions to their usual activities due to stomach pain or bowel problems or unintentional weight loss.

Pre-survey questions survey addressing healthcare provider recommendations, perceived risk of gastric cancer, and reasons for undergoing the current colonoscopy were measured by binary (yes/no) and multiple-choice questions.

*Perceptions and Beliefs About Gastric Cancer Screening- Pre-EGD survey*

Perceptions and beliefs about gastric cancer screening was measured using several domains, including perceived susceptibility, severity, benefits, efficacy, and barriers to gastric cancer screening, fatalism, destiny, and trust in the health care system were assessed using Likert-type items, ranging from 1 (strongly disagree) to 5 (strongly agree). Perceived threat was evaluated combining perceived susceptibility and severity. Perceived susceptibility captured participants' beliefs about their personal risk of developing gastric cancer based on factors such as family history, symptoms, or prior medical conditions. Perceived severity addressed beliefs about the seriousness and potential health consequences of gastric cancer. Fatalism and destiny constructs measured beliefs regarding the inevitability of cancer and the perceived influence of individual actions on health outcomes. Perceived benefits assessed participants' views on the value of gastric cancer screening, including early detection of cancer or precancerous lesions and improved treatment outcomes. Self-efficacy measured participants' confidence in their ability to complete the recommended screening procedures. Perceived barriers encompassed logistical, financial, emotional, and access-related obstacles that might impede screening. Health care access barriers included difficulty obtaining transportation and difficulty understanding written medical information. Trust in the health care system was evaluated through statements assessing perceived quality of care, intentions of the system, and concerns regarding unethical practices. These constructs were analyzed descriptively and retained at the item level with higher scores indicating stronger agreement with each corresponding belief domain. A composite score was calculated for each domain.

*Attitudes and Perceptions Toward Opportunistic EGD- Post-EGD survey*

Participant-reported perceptions related to gastric cancer screening after EGD procedure were assessed in a post-EGD procedure survey including 24 questions. Overall satisfaction was measured with satisfaction with having an EGD at the same time of colonoscopy and the perceived easiness of completing them. Preference items measured respondents' desired timing and sequencing of EGD relative to screening colonoscopy, willingness to undergo combined or separate procedures, inclination to request EGD in clinical encounters, and likelihood of recommending combined EGD and colonoscopy to others. Perceived efficacy was assessed through items evaluating the ease, convenience, and perceived informational value of EGD for understanding stomach cancer. Screening barrier and risk-assessment items measured competing priorities and practical constraints, including caregiving demands and overall prioritization of stomach cancer screening. Screening acceptability was evaluated using an established multidimensional framework that assessed affective attitude (comfort with undergoing EGD), perceived burden (effort required to participate), ethicality (alignment with personal values), perceived effectiveness (expected impact on stomach cancer outcomes), intervention coherence (understanding of EGD's purpose in detection or management), self-efficacy (confidence in completing future EGDs), opportunity cost (extent to which undergoing EGD interferes with other priorities), and overall acceptability. Responses were collected using Likert-type scales to quantify the extent of agreement, willingness, or behavioral inclination across these domains.

*Procedures and Biospecimen Collection*

Urine and saliva samples were collected before the start of the EGD. Stool samples were collected using kits mailed to participants and returned to the facility for processing. Blood samples were obtained during routine intravenous placement prior to the EGD/colonoscopy procedure. All biological samples were processed, aliquoted, and stored at –80°C until further analysis.

EGD and Biopsy Procedure: During EGD, biopsies were obtained from standard gastric sites: the lesser and greater curvatures of the antrum, the lesser and greater curvatures of the body, and the incisura angularis. Evaluation for pre-cancerous lesions followed the updated Sydney System, the international standard for gastritis classification. The Operative Link on Gastric Atrophy (OLGA) and Operative Link on Gastric Intestinal Metaplasia (OLGIM) systems were used to assess the severity and distribution of atrophic gastritis (AG) and gastric intestinal metaplasia (IM). IM, when present, was subtyped as complete, incomplete, or mixed using standard Hematoxylin and Eosin (H&E) staining. Dysplasia was scored according to the Vienna system, and detection of active *Helicobacter pylori* infection using H&E and immunohistochemical staining.

**Data Management and Coding**

All variables underwent quality checks for completeness, range validity, and internal consistency. Sociodemographic variables were categorized to preserve interpretability. Perceptions and attitudes were coded directionally (e.g., higher values indicating greater agreement or greater perceived barriers). Missing values were handled using available-case analysis, unless that information can be found in the medical record. Implausible entries were reviewed and corrected where possible; otherwise, they were recorded as missing. Binary, ordinal, and continuous variables were prepared according to the requirements of planned statistical analyses.

### Supplementary Table 1. Pre- and Post EGD Survey items

| Pre-survey Items | Domain | Source |
|---|---|---|
| Diet (18 questions) | Pickled food | ACS Cancer Prevention Study Questionnaire[1] |
| | Non-fermented soy food | |
| | Food cooked with garlic, onion, chives | |
| | Salt added at the table | |
| | Spicy food or added hot spices | |
| | Fruit and vegetable intake | |
| | Processed meats | |
| | Dairy products | |
| | Diet habits | |
| Lifestyle (7 questions) | Moderate and vigorous physical activity | ACS Cancer Prevention Study Questionnaire[1] |
| | Tobacco smoking, | |
| | Alcohol consumption per day | |
| Medical History (10 questions) | Anthropometry | NIH-AARP study Questionnaire 1995[2] |
| | Comorbidities and surgery history | Developed by the study team |
| | Infection treatment history | Developed by the study team |
| | Blood type | Developed by the study team |
| | Family history of gastric cancer | Based on Dhillon 2001 study[3] |
| | Medication history | NIH-AARP Risk Factor Questionnaire 1995[2] |
| | Current symptoms | Developed by the study team |
| Health beliefs (28 questions) | Perceived susceptibility | Risk Behavior Diagnosis Scale (RBDS)[4-6]<br>Fear of GC: Affect in Risk Scale[5-9] |
| | Perceived severity | |
| | Response efficacy | |
| | Benefits | |
| Screening barriers (26 questions) | Competing life concerns | Competing life concerns[5, 6, 10] |
| | Fatalism and Destiny | Fatalism and Destiny[5, 6, 11, 12] |
| | Health care distrust | Health System Distrust Scale[5, 13] |
| | Screening facilitators and barriers | Facilitators and Barriers[5, 6, 10, 14-16] |
| Gastric Cancer Knowledge (20 questions) | Gastric Cancer risk factors | Developed by the study team |
| Perceived risk/ provider recommendation (3 questions) | Test Accuracy and preferences | Gastrointestinal Endoscopy Satisfaction Questionnaire (GESQ)[17] |
| Colonoscopy outcomes (5 questions) | Colonoscopy history, perceptions, and preferences | Colonoscopy preferences[5, 18] |
| Health literacy and language (8 questions) | Reading literacy, writing literacy, childhood language, social language, thinking language | Health Literacy[19, 20] |
| Culture and Background (9 questions) | Country of origin | HCHS/SOL Personal information questionnaire[21] |
| | Immigration history | Cha-Cha Demographic Questionnaire |
| | Acculturation | Short Acculturation Scale for Hispanics (SASH)[22, 23] |

| | | |
|---|---|---|
| | Ethnic food intake | Developed by the study team |
| | Education | NIH-AARP study Questionnaire 1995[2] |
| | Race/ethnicity | U.S census bureau American community survey[24] |
| | Insurance type | HCHS/SOL Economic Questionnaire[21] |
| **Post-survey Items** | **Domain** | **Source** |
| Satisfaction (2 questions) | Satisfaction, easiness | Gastrointestinal Endoscopy Satisfaction Questionnaire (GESQ)[17] |
| Preference (6 questions) | Preferences | Colonoscopy preferences[5, 18] Developed by the study team |
| Perceived risk/ provider recommendation (1 question) | Test accuracy and preferences | Gastrointestinal Endoscopy Satisfaction Questionnaire (GESQ)[17] |
| Risk assessment /Screening (8 questions) | Risk assessment | Gastrointestinal Endoscopy Satisfaction Questionnaire (GESQ)[17] |
| Acceptability (8 questions) | Affective attitude, procedure burden, ethical concerns, perceived effectiveness, intervention coherence, self-efficacy, opportunity cost, and overall acceptability | Based on Sekhon's 2022 study[25] |

### Supplementary Table 2. Reasons for refusal

| High Risk (n=36) 34.3% | Low Risk (n=55) 66.3% |
|---|---|
| Not interested in endoscopy (n=16) | Not interested in endoscopy (n=23) |
| Rescheduling (n=11) | Rescheduling at an unforeseen date (n =12) |
| Fear of additional anesthesia (n=6) | Fear of additional anesthesia (n=9) |
| Having no time (n=2) | Fear of having biopsies taken (n=4) |
| Overwhelmed by process (n=1) | Having no time (n=2) |
| Did not want to be a part of research (n=1) | Wasn't informed by their doctor (n=2) |
| | Privacy concern (n=1) |
| | Colonoscopy scheduled outside the institution (n=1) |
| | Fear of endoscopy (n=1) |

Supplementary Table 3. Post-survey on the perception of gastric cancer screening using a combined Endoscopy and Colonoscopy procedure by risk groups (n=40)

| | | High-risk (n=27) | Low-risk (n=13) | p-value |
|---|---|---|---|---|
| **Satisfaction with Colonoscopy and Endoscopy Outcomes** | | | | |
| | 1. Satisfaction with having EGD at the same time as the colonoscopy | | | |
| | • Very satisfied/satisfied<br>• Neutral<br>• Dissatisfied/very dissatisfied | 100.0<br>0.0<br>0.0 | 100.0<br>0.0<br>0.0 | 0.0776 |
| | 2. Easy or difficult to have EGD at the same time as the colonoscopy | | | |
| | • Very easy/Easy<br>• No difference<br>• Difficult/very difficult | 92.6<br>7.4<br>0.0 | 100.0<br>0.0<br>0.0 | 0.4335 |
| **Preference for the addition of EGD and its timing and sequence** | | | | |
| | 1. Preference for getting upper endoscopy (EGD) | | | |
| | • At the same time as the screening colonoscopy<br>• At a separate time from the screening colonoscopy | 100.0<br>0.0 | 92.3<br>7.7 | 0.3250 |
| | 2. Inclination to undergo screening with the addition of EGD | | | |
| | • Much more/somewhat<br>• Not change whether I want to undergo screening<br>• Somewhat less/ much less | 88.9<br>7.4<br>3.7 | 61.5<br>38.5<br>0.0 | 0.3327 |
| | 3. Willingness to get both upper endoscopy (EGD) and colonoscopy at the same time in the future (if the doctor recommends EGD-SC combined) | | | |
| | • Very willing/Willing<br>• Neutral<br>• Unwilling/very unwilling | 96.3<br>3.7<br>0.0 | 100.0<br>0.0<br>0.0 | 0.2665 |
| | 4. Willingness to undergo upper endoscopy (EGD) as a separate procedure in the future, if the doctor recommends EGD separately from SC) | | | |
| | • Very willing/Willing<br>• Neutral<br>• Unwilling/very unwilling | 51.9<br>22.2<br>25.9 | 61.5<br>23.1<br>15.4 | 0.3087 |
| | 5. Inclination to ask to add an EGD to an SC, if the doctor recommends SC without mentioning EGD | | | |
| | • Inclined/strongly inclined<br>• Inclined but would not insist if they did not recommend it<br>• Not inclined to ask unless I was asked first | 74.1<br>18.5<br>7.4 | 69.2<br>15.4<br>15.4 | 0.8367 |
| | 6. Recommendation of combined EGD+SC to family or friends | | | |
| | • Very likely/Likely<br>• Neutral<br>• Not likely/Very unlikely | 92.6<br>7.4<br>0.0 | 92.3<br>7.7<br>0.0 | 0.2540 |
| **Efficacy** | | | | |
| | 1. Easiness of having an EGD | | | |
| | • Strongly agree/Agree<br>• Neither disagree nor agree<br>• Disagree/Strongly disagree<br>• No answer | 40.8<br>37.0<br>14.8<br>7.4 | 30.8<br>53.8<br>15.4<br>0.0 | 0.7202 |

| | | | | |
|---|---|---|---|---|
| | 2. Learn more about stomach cancer with an EGD | | | |
| | • Strongly agree/Agree<br>• Neither disagree nor agree<br>• Disagree/Strongly disagree | 81.5<br>14.8<br>3.7 | 92.3<br>7.7<br>0.0 | 0.0066 |
| | 3. Convenience of having an EGD | | | |
| | • Strongly agree/Agree<br>• Neither disagree nor agree<br>• Disagree/Strongly disagree<br>• No answer | 48.2<br>40.7<br>3.7<br>7.4 | 38.5<br>61.5<br>0.0<br>0.0 | 0.7475 |
| **Risk Assessment/ Screening barriers** | | | | |
| | 1. Too many other things to worry about compared to getting stomach cancer screening | | | |
| | • Strongly agree/Agree<br>• Neither disagree nor agree<br>• Disagree/Strongly disagree | 18.5<br>11.1<br>70.4 | 0.0<br>0.0<br>100.0 | 0.0656 |
| | 2. Stomach cancer screening is a low priority on my list | | | |
| | • Strongly agree/Agree<br>• Neither disagree nor agree<br>• Disagree/Strongly disagree | 3.7<br>11.1<br>85.2 | 7.7<br>7.7<br>84.6 | 0.8859 |
| | 3. Too busy taking care of my children or other family members to get stomach cancer screening at this time. | | | |
| | • Strongly agree/Agree<br>• Neither disagree nor agree<br>• Disagree/Strongly disagree | 11.1<br>11.1<br>77.8 | 0.0<br>7.7<br>92.3 | 0.0515 |
| **Acceptability** | | | | |
| | 1. Feels comfortable getting an EGD (affective attitude) | | | |
| | • Very comfortable/Comfortable<br>• No opinion<br>• Uncomfortable/very uncomfortable | 70.4<br>18.5<br>11.1 | 84.6<br>15.4<br>0.0 | 0.0427 |
| | 2. The amount of effort required to participate in the EGD (burden) | | | |
| | • No effort at all/ a little effort<br>• No opinion<br>• A lot of effort/Huge effort | 96.3<br>0.0<br>3.7 | 92.3<br>7.7<br>0.0 | 0.4477 |
| | 3. Individual's value system about getting an EGD (Ethicality) | | | |
| | • Very moral/Moral<br>• No opinion<br>• Unmoral/ Very unmoral | 63.0<br>25.9<br>11.1 | 76.9<br>15.4<br>7.7 | 0.5271 |
| | 4. The potential of EGD to positively impact outcomes related to stomach cancer (perceived effectiveness) | | | |
| | • Strongly agree/Agree<br>• Neither disagree nor agree<br>• Disagree/Strongly disagree | 70.4<br>22.2<br>7.4 | 100.0<br>0.0<br>0.0 | 0.0246 |
| | 5. Understands how EGD helps to detect and/or manage stomach cancer (intervention coherence) | | | |
| | • Strongly agree/Agree<br>• Neither disagree nor agree<br>• Disagree/Strongly disagree | 81.5<br>11.1<br>7.4 | 100.0<br>0.0<br>0.0 | 0.4016 |
| | 6. Confidence about getting an EGD in the future (self-efficacy) | | | |
| | • Very confident/confident<br>• No opinion<br>• Unconfident/Very unconfident | 85.2<br>7.4<br>7.4 | 100.0<br>0.0<br>0.0 | 0.6026 |

| | | | | |
|---|---|---|---|---|
| | 7. Interference of getting an EGD with other priorities (opportunity cost) | | | |
| | • Strongly agree/Agree<br>• Neither disagree nor agree<br>• Disagree/Strongly disagree | 7.4<br>3.7<br>88.9 | 0.0<br>0.0<br>100.0 | 0.0050 |
| | 8. Acceptability of getting an EGD (general acceptability) | | | |
| | • Completely acceptable/Acceptable<br>• No opinion<br>• Unacceptable/Completely unacceptable | 92.6<br>0.0<br>7.4 | 84.6<br>7.7<br>7.7 | 0.6453 |

Supplementary Table 4. Baseline screening behaviors (n=43)

| **Colonoscopy Screening** | **N (%)** |
|---|---|
| 1. Prior colonoscopy (Yes) | 24 (55.8) |
| 2. Reason for scheduling current colonoscopy (more than one answer per participant) | |
| - Desire to know if I have cancer | 8 (18.6) |
| - Reduce fear and concerns about colon cancer | 14 (32.6) |
| - Someone in my family has/had cancer | 7 (16.3) |
| - Colonoscopy seems important | 15 (34.9) |
| - Family members wanted me to go | 3 (7.0) |
| - Saw/read about colonoscopy in the media | 0 (0.0) |
| - My doctor recommended that I go | 27 (62.8) |
| - Desire to know if my colon is healthy | 17 (39.5) |
| - Others (time for screening, positive fecal test, symptoms, family has polyps) | 8 (18.6) |
| 3. Reason for current colonoscopy visit | |
| - Follow-up for polyps on my prior colonoscopy | 9 (20.9) |
| - Screening for colon cancer | 25 (58.1) |
| - Screening for non-cancerous diseases | 6 (14.0) |
| - A prior test showed a positive cancer result | 1 (2.3) |
| - Diagnostic - I have potential cancer symptoms | 0 (0.0) |
| - Diagnostic - I have gastrointestinal disease symptoms | 2 (4.7) |
| 4. Feelings about having colonoscopy for personal health and well-being | |
| - Very critical/ Critical | 38 (88.4) |
| - Neither critical nor uncritical | 5 (11.6) |
| - Uncritical/Very uncritical | 0 (0.0) |
| 5. Reasons for electing screening colonoscopy as opposed to other options (more than one answer) | |
| - Colonoscopy would be more effective than the mail-in fecal test | 11 (25.6) |
| - Colonoscopy results would be more reliable than the mail-in fecal test | 13 (30.2) |
| - Colonoscopy would be better for screening and/or diagnosis | 18 (41.9) |
| - Not given alternative options other than colonoscopy | 5 (11.6) |
| - Family members wanted me to go for a colonoscopy | 2 (4.7) |
| - Doctor recommendation for a colonoscopy | 26 (60.5) |
| - Personal choice to go for a colonoscopy | 13 (30.2) |
| - Desire to know future risk of cancer | 14 (32.6) |
| - Colonoscopy would be easier the mail-in fecal test | 1 (2.3) |
| - Colonoscopy would be less expensive than the mail-in fecal | 0 (0.0) |
| - Colonoscopy was covered by my insurance | 10 (23.3) |
| - Colonoscopy would reduce fear and concerns about cancer | 13 (30.2) |
| - Someone in their family has a colon cancer | 6 (14.0) |
| - Going for a colonoscopy seemed important | 18 (41.9) |
| - Others (age, constipation, family member has polyps) | 3 (7.0) |
| **Gastric cancer perceived risk and provider recommendations beliefs** | |
| 1 Discussed Personal GC risk with healthcare provider (Yes) | 6 (14.0) |
| 2. Healthcare provider ever recommended getting screening for stomach cancer (Yes) | 8 (19.5) |

Supplementary Table 5. Perceptions and beliefs of susceptibility, severity benefit and barriers about gastric cancer screening by high and low risk groups (n=43)

| **Perceived:** | **Mean score, (SD) (1-5 scale)** | | **Agree/ Strongly Agree, (≥4) N (%)** | | **Disagree/ Strongly Disagree (≤2) N (%)** | | **Neither agree/ disagree, (3) (N%)** | | **p-value[b]** |
|---|---|---|---|---|---|---|---|---|---|
| | **High-risk** | **Low-risk** | **High-risk** | **Low-risk** | **High-risk** | **Low-risk** | **High-risk** | **Low-risk** | |
| **Susceptibility** | 2.6 (0.7) | 2.3 (0.9) | 0 (0.0) | 0 (0.0) | 7 (25.9) | 7 (43.8) | 20 (74.1) | 9 (56.3) | 0.2451 |
| **Severity** | 3.5 (0.7) | 3.5 (0.6) | 6 (22.2) | 4 (25.0) | 1 (3.7) | 0 (0.0) | 20 (74.1) | 12 (75.0) | 0.9900 |
| **Barriers** | 2.1 (0.6) | 1.9 (0.6) | 0 (0.0) | 0 (0.0) | 12 (44.4) | 12 (75.0) | 15 (55.6) | 4 (25.0) | 0.0757 |
| **Benefits** | 4.0 (0.7) | 4.3 (0.5) | 18 (66.7) | 13 (81.3) | 1 (3.7) | 0 (0.0) | 8 (29.6) | 3 (18.7) | 0.0628 |

[a] 43 respondents completed the pre-EGD survey
[b] P-values were calculated based on Mann-Whitney U test to examine the differences in survey scale responses between high- and low-risk participants

Supplementary Table 6. Thoughts about screening before and after the combined EGD-SC procedure.

| **Thoughts about talking to a healthcare provider about EGD for stomach cancer screening in the future?** | Pre-survey (n=43) | Post-survey (n=40) |
|---|---|---|
| • I do not want to think about stomach cancer screening right now | 5 (11.6) | 2 (4.9) |
| • I have thought about it and am undecided about whether to talk with a healthcare provider about having stomach cancer screening | 6 (14.0) | 9 (22.0) |
| • I have thought about it, but I do not want to talk with a healthcare provider about stomach cancer screening | 2 (4.7) | 1 (2.4) |
| • I have decided I would like to talk with a healthcare provider about stomach cancer screening, but I don't know when | 9 (20.9) | 7 (17.1) |
| • I will definitely talk to a healthcare provider about stomach cancer screening in the next six months/next available opportunity | 14 (32.6) | 19 (46.3) |
| • I have already talked with a healthcare provider about stomach cancer screening | 7 (16.3) | 3 (7.32) |

Supplementary Table 7. Gastrointestinal symptoms and pathological findings

| Pathologic findings[a] | Symptomatic | No GI symptoms | P-value |
| --- | --- | --- | --- |
| AG | 4 (14.8) | 2 (12.5) | 1.000 |
| IM | 3 (11.1) | 1 (6.25) | 1.000 |
| HP | 9 (33.3) | 6 (37.5) | 1.000 |
| AG, IM, or dysplasia | 5 (18.5) | 2 (23.5) | 0.695 |
| HP, AG, IM or dysplasia | 12 (44.4) | 6 (37.5) | 0.755 |

Abbreviations: AG, atrophic gastritis; IM, intestinal metaplasia; HP, Helicobacter pylori
[a] Based on 43 respondents who completed the pre-EGD survey
Symptomatic individuals were defined as those who reported experiencing stomach pain or nausea at least once per week; vomiting at least once per month; frequent bloating; changes in appetite; or interruptions to their usual activities due to stomach pain or bowel problems or unintentional weight loss.

Supplementary Table 8. Clinically reported findings from EGD

| | Clinical findings (n=50) |
| --- | --- |
| **Gastric endoscopy clinical report** | |
| Polyps | 10 (20.0) |
| Gastritis | 33 (66.0) |
| **Gastric pathological-clinical report (31 biopsies)** | |
| Gastritis | 14 (28.0) |
| Atrophic Gastritis | 1 (2.0) |
| Intestinal Metaplasia | 2 (4.0) |
| *H. pylori* diagnosis (post EGD) | 10 (20.0) |
| **Esophageal endoscopy clinical report** | |
| Normal findings | 39 (78.0) |
| Mild esophagitis (LA Grade A-B) | 7 (14.0) |
| Hiatus hernia only (Hill Grade III-IV) | 1 (2.0) |
| Severe esophagitis (LA Grade C-D) | 1 (2.0) |
| Single nodule | 1 (2.0) |
| Esophageal plaques | 1 (2.0) |
| **Esophageal Pathological-clinical report (2 biopsies)** | |
| Squamous papilloma | 1 (2.0) |
| Esophageal candidiasis | 1 (2.0) |

Abbreviations: LA Grade; Los Angeles classification system for reflux esophagitis

**Supplementary Table 1 References**

11. Kobayashi LC, Smith SG. Cancer Fatalism, Literacy, and Cancer Information Seeking in the American Public. Health Educ Behav. 2016;43(4):461-70. Epub 20150916. doi: 10.1177/1090198115604616. PubMed PMID: 26377524; PMCID: PMC5123630.
12. Niederdeppe J, Levy AG. Fatalistic beliefs about cancer prevention and three prevention behaviors. Cancer Epidemiol Biomarkers Prev. 2007;16(5):998-1003. doi: 10.1158/1055-9965.EPI-06-0608. PubMed PMID: 17507628.
13. Shea JA, Micco E, Dean LT, McMurphy S, Schwartz JS, Armstrong K. Development of a revised Health Care System Distrust scale. J Gen Intern Med. 2008;23(6):727-32. Epub 20080328. doi: 10.1007/s11606-008-0575-3. PubMed PMID: 18369678; PMCID: PMC2517896.
14. Anderson B, McLosky J, Wasilevich E, Lyon-Callo S, Duquette D, Copeland G. Barriers and facilitators for utilization of genetic counseling and risk assessment services in young female breast cancer survivors. J Cancer Epidemiol. 2012;2012:298745. Epub 20121022. doi: 10.1155/2012/298745. PubMed PMID: 23150731; PMCID: PMC3485517.
15. Gammon AD, Rothwell E, Simmons R, Lowery JT, Ballinger L, Hill DA, Boucher KM, Kinney AY. Awareness and preferences regarding BRCA1/2 genetic counseling and testing among Latinas and non-Latina white women at increased risk for hereditary breast and ovarian cancer. J Genet Couns. 2011;20(6):625-38. Epub 20110621. doi: 10.1007/s10897-011-9376-7. PubMed PMID: 21691939; PMCID: PMC7373795.
16. Kinney AY, Gammon A, Coxworth J, Simonsen SE, Arce-Laretta M. Exploring attitudes, beliefs, and communication preferences of Latino community members regarding BRCA1/2 mutation testing and preventive strategies. Genet Med. 2010;12(2):105-15. doi: 10.1097/GIM.0b013e3181c9af2d. PubMed PMID: 20061960; PMCID: PMC3022322.
17. Hutchings HA, Cheung WY, Alrubaiy L, Durai D, Russell IT, Williams JG. Development and validation of the Gastrointestinal Endoscopy Satisfaction Questionnaire (GESQ). Endoscopy. 2015;47(12):1137-43. Epub 20150908. doi: 10.1055/s-0034-1392547. PubMed PMID: 26349066.
18. Bie AKL, Brodersen J. Why do some participants in colorectal cancer screening choose not to undergo colonoscopy following a positive test result? A qualitative study. Scand J Prim Health Care. 2018;36(3):262-71. doi: 10.1080/02813432.2018.1487520. PubMed PMID: 30238859; PMCID: PMC6381546.
19. Chew LD, Bradley KA, Boyko EJ. Brief questions to identify patients with inadequate health literacy. Fam Med. 2004;36(8):588-94. PubMed PMID: 15343421.
20. Chew LD, Griffin JM, Partin MR, Noorbaloochi S, Grill JP, Snyder A, Bradley KA, Nugent SM, Baines AD, Vanryn M. Validation of screening questions for limited health literacy in a large VA outpatient population. J Gen Intern Med. 2008;23(5):561-6. Epub 20080312. doi: 10.1007/s11606-008-0520-5. PubMed PMID: 18335281; PMCID: PMC2324160.
21. National Heart L, and Blood Institute. Hispanic Community Health Study/Study of Latinos (HCHS/SOL) National Heart, Lung, and Blood Institute2025 [cited 2025 December 2025]. Available from: https://www.nhlbi.nih.gov/science/hispanic-community-health-studystudy-latinos-hchssol.
22. Hamilton AS, Hofer TP, Hawley ST, Morrell D, Leventhal M, Deapen D, Salem B, Katz SJ. Latinas and breast cancer outcomes: population-based sampling, ethnic identity, and

acculturation assessment. Cancer Epidemiol Biomarkers Prev. 2009;18(7):2022-9. Epub 20090623. doi: 10.1158/1055-9965.EPI-09-0238. PubMed PMID: 19549806; PMCID: PMC4147726.
23. Marin G, Sabogal F, Marin BV, Oterosabogal R, Perezstable EJ. Development of a Short Acculturation Scale for Hispanics. Hispanic J Behav Sci. 1987;9(2):183-205. doi: Doi 10.1177/07399863870092005. PubMed PMID: WOS:A1987K299900005.
24. Bureau USC. American Community Survey (ACS): United States Census Bureau; [cited 2025 December 2025]. Available from: https://www.census.gov/programs-surveys/acs.html.
25. Sekhon M, Cartwright M, Francis JJ. Development of a theory-informed questionnaire to assess the acceptability of healthcare interventions. BMC Health Serv Res. 2022;22(1):279. Epub 20220301. doi: 10.1186/s12913-022-07577-3. PubMed PMID: 35232455; PMCID: PMC8887649.
